# Antibody Persistence and Safety through 6 Months after Heterologous Orally Aerosolised Ad5-nCoV in individuals primed with two-dose CoronaVac previously

**DOI:** 10.1101/2022.07.26.22278072

**Authors:** Lairun Jin, Rong Tang, Shipo Wu, Xiling Guo, Lihua Hou, Xiaoqin Chen, Tao Zhu, Jinbo Gou, Haitao Huang, Jin Zhong, Hongxing Pan, Lunbiao Cui, Yin chen, Xin Xia, Jialu Feng, Xue Wang, Qi Zhao, XiaoYu Xu, Zhuopei Li, Xiaoyin Zhang, Jingxin Li, Wei Chen, Fengcai Zhu

## Abstract

**Background:** Heterologous orally administered adenovirus type-5 vector-based COVID-19 vaccine (Ad5-nCoV) in individuals who were primed with two-dose CoronaVac (an inactivated SARS-CoV-2 vaccine, by Sinovac) previously, has been reported to be safe and highly immunogenic within 28 days post-boosting. However, antibody persistence and safety up to 6 months of this regimen are not been reported yet.

**Methods:** This is a randomized, open label, single-center trial on safety and immunogenicity of heterologous boost immunization with an orally administered aerosolised Ad5-nCoV vs. homologous boost immunization with CoronaVac after two-dose priming with CoronaVac in Chinese adults aged 18 years and older (NCT05043259). We followed the participants in this trial, including 140 in the low-dose aerosolised Ad5-nCoV group, 139 in the high-dose aerosolised Ad5-nCoV group, and 140 in the CoronaVac group for 6 months. Neutralising antibodies (NAbs) against live wild-type SARS-CoV-2 virus and omicron variant, and receptor-binding domain (RBD)-specific IgG antibodies were detected in serum samples collected at 28 days, 3 months, and 6 months after the booster dose. Serious adverse events (SAEs) were documented till month 6.

**Results:** The low-dose and high-dose heterologous boost immunisation groups had NAb GMTs against live wild-type SARS-CoV-2 of 1937.3 [95% CI 1466.9, 2558.4] and 1350.8 [95% CI 952.6, 1915.3], which were 26.4 folds and 18.4 folds higher than that the CoronaVac group did (73.5 [95% CI 52.3, 103.3]) at 28 days. The low-dose and high-dose heterologous boost immunisation groups had NAb GMTs against live wild-type SARS-CoV-2 of 530.1 (95% CI 412.5, 681.1) and 457.6 (95%CI 349.4, 599.2), which were 26.0 folds and 22.4 folds higher than that the CoronaVac group did (20.4 [95%CI 14.3, 29.1]) at 3 months, respectively. At 6 months, the low-dose and high-dose heterologous booster groups had NAb GMTs against live wild-type SARS-CoV-2 of 312.9 (95% CI 237.7, 411.8) and 251.1 (95% CI 178.2, 354.0), which were 30.1 folds and 24.1 folds higher than the CoronaVac group did (10.4 [95% CI 7.8, 14.0]), respectively. Additionally, the low-dose and high-dose heterologous booster groups had NAb GMTs against live omicron variant of 52.0 (95% CI 37.2, 72.6) and 23.1 (95% CI 15.7, 33.9) at 28 days, 27.9 (95% CI 18.8, 41.3) and 23.3 (95% CI 16.2, 33.3) at 3 months, 16.0 (95% CI 10.9, 23.5) and 12.0 (95% CI 8.5, 16.8) at 6 months, respectively. However, nearly all participants had no detectable NAbs for omicron variant in the CoronaVac group at either 28 days, 3 months, or 6 months. No vaccine-related SAEs were observed.

**Conclusions:** These data suggested that heterologous aerosolised Ad5-nCoV following two-dose CoronaVac priming was safe and persistently more immunogenic than three-dose CoronaVac, although immune responses waned over time.

## Introduction

The COVID-19 pandemic caused by SARS-CoV-2 is profoundly affecting life around the world. Globally, as of 14 June 2022, there have been more than 0.5 billion confirmed cases of COVID-19, including nearly 6.31 million deaths, and the number of cases continues to rise [1]. Safe and effective vaccination strategies are urgently needed to control this pandemic. Global mass vaccination has played an effective role in reducing SARS-CoV-2 transmission, hospitalizations and deaths. As of June 14, 2022, 11 vaccines have been granted emergency use by WHO, all of which were administered intramuscularly [2].

The emergence and rapid spread of SARS-CoV-2 variants of concern [3] and waning immunity over time [4] are challenging the effectiveness of vaccines. In view of this emerging evidence, the WHO Strategic Advisory Group of Experts on Immunization recommends that booster (third) doses should be offered 4-6 months after completion of the primary vaccination series, especially for inactivated vaccines. Both homologous (same vaccine platform) and heterologous (different vaccine platform) vaccines could be used for such booster doses [5].

CoronaVac, the Sinovac inactivated whole-virion SARS-CoV-2 vaccine, has been widely used in a two-dose regimen as primary series. Previous studies have shown that both homologous booster with CoronaVac and heterologous booster with a recombinant adenoviral-vectored ChAdOx1 nCoV-19 vaccine (AZD1222, AstraZeneca) induced more robust immune responses than primary immunization of two-dose CoronaVac [6, 7]. Two randomized controlled trials of adults who had received two doses of CoronaVac showed that all four heterologous regimens (an RBD-subunit vaccine, ZF2001, Anhui Zhifei Longcom; a recombinant adenoviral vectored vaccine, Ad26.COV2-S, Janssen; an mRNA vaccine, BNT162b2, Pfizer–BioNTech; or AZD1222, respectively, as a booster dose) could enhance immunogenicity after the booster dose, compared with a third homologous dose of CoronaVac [8, 9]. A large-scale prospective cohort study of individuals who completed their primary series with CoronaVac showed that heterologous booster immunization with AZD1222 or BNT162b2 provided higher vaccine effectiveness than a homologous CoronaVac booster for all COVID-19 outcomes, including symptomatic COVID-19, severe disease and death [10]. These studies supported a heterologous booster regimen following completion of two-dose CoronaVac. However, these results were limited to 28 days or 3 months after the booster dose. The long-lasting protection of heterologous prime-boost regimens is currently unknown.

In our previously reported study [11], a heterologous booster vaccine with an orally administered aerosolised Ad5-nCoV (by CanSino Biologics, China) induced substantial serum NAb titers against SARS-CoV-2, which were 6.7-10.7-fold stronger serum NAbs response than a homologous CoronaVac booster between days 14 and 28 after the booster dose. Participants who inhaled 0.1 mL and 0.2 mL of aerosolized Ad5-nCoV had NAb GMT peaks of 6054.1 and 4221.3 IU/mL against live SARS-CoV-2 virus, respectively, at day 28 after the booster vaccination, which was higher than three doses of BNT162b2 (955.7 IU/mL). In addition, a heterologous booster vaccine with an orally administered aerosolised Ad5-nCoV was safe and had lower adverse reactions than a homologous boost with CoronaVac in adults who have previously received two-dose CoronaVac. However, antibody persistence and long-term safety of this vaccination regimen are not been reported yet. Here, we updated the serological and safety data to 6 months from participants following the heterologous orally administration with aerosolised Ad5-nCoV in Chinese adults or a homologous immunization with CoronaVac in this trial.

## Methods

### Study design and samples

We did a randomised, open label, single-centre trial on safety and immunogenicity of heterologous boost immunisation with an orally administered aerosolised Ad5-nCoV after two-dose priming with an inactivated SARS-CoV-2 vaccine in Chinese adults aged 18 years and older (NCT05043259), which has been reported previously. Briefly, healthy participants who had received two-dose CoronaVac as primary immunization (3 to 9 months after dose 2) were recruited from Lianyungang, Jiangsu, China. SARS-CoV-2 infection or laboratory-confirmed COVID-19, nasal or oral disease, psychosis, abnormal lung function, severe cardiovascular disease, HIV infection and positive urine pregnancy were excluded. 140 participants received a heterologous booster immunization with aerosolised Ad5-nCoV at 0.1mL in the low-dose group, 139 received a heterologous booster immunization with aerosolised Ad5-nCoV at 0.2mL in the high-dose group, and 140 received a homologous intramuscular booster immunization in the CoronaVac group. Written informed consent was obtained from all participants. The trial protocol was approved by Research Ethics Committee of Jiangsu Provincial Center for Disease Control and Prevention, and was conducted following the principles of the Declaration of Helsinki, ICH Good Clinical Practice guidelines and local guidelines.

We have previously reported the incidence of adverse reactions within 14 days post-boost, serious adverse events during the first 28 days post-boost vaccination, the serum NAbs against live SARS-CoV-2 virus and delta variants, receptor-binding domain (RBD)-specific binding IgG and IgA responses at days 14 and 28 after the booster dose, and SARS-CoV-2 spike-specific cytokine T cells responses after receiving a boost vaccination [11]. In this report, we revealed the neutralising antibodies (NAbs) against live wild-type SARS-CoV-2 and omicron variant and RBD-specific IgG antibodies in serum of the first 40 participants in each of the three treatment groups at months 3 and 6 after the booster dose. We also reported the serious adverse events (SAEs) occurred through 6 months after booster vaccination and pregnancy events.

### Serologic Assays

Serum samples were collected from 120 participants (first 40 participants in each group) at months 3 and 6 for antibody persistency analysis. NAb titers against the live wild-type SARS-CoV-2 and the omicron (B.1.1.529) variant were determined by using cytopathic effect-based microneutralisation assay with a wild-type SARS-CoV-2 virus isolate BetaCoV/Jiangsu/JS02/2020 (GISAID EPI_ISL_411952) and an omicron (B.1.1.529) variant isolate hCoV-19/Jiangsu/JS01/2022 (GISAID EPI_ISL_12511653) (BA.1) in Vero-E6 cells (National Collection of Authenticated Cell Cultures, National Academy of Science, China). The serum dilution for microneutralisation assay was 1:8 to 1:8192, and then mixed with the equal volume of virus solution to reach a 50% tissue culture infectious dose of 100 per well. The reported titers were the reciprocal of the highest sample dilution observed by an inverted microscope protecting at least 50% of the cells from cytopathy. The seropositivity for NAbs was defined as titer ≥1:8. We used the WHO International Standard (NIBSC code 20/136) as the reference for the cytopathic effect-based microneutralisation assays. We used the commercial Anti-SARS-CoV-2 RBD IgG ELISA kit (Vazyme Medical Technology, Nanjing, China) to measure RBD-specific IgG responses (RU/ml) with a cutoff titer of 1:10 [11].

### Statistical analysis

The sample size calculation was based on the hypothesis that a booster vaccination with aerosolised Ad5-nCoV following the two doses of CoronaVac could elicit a non-inferior or superior serum NAbs response at 14 days after the booster dose compared with a third dose of CoronaVac does, which has been reported previously [11]. Serum NAbs titres against the wild-type SARS-CoV-2 virus and omicron (B.1.1.529) variant at day 28, month 3, and month 6 were measured in a subgroup of the first 40 participants out of 150 participants in each group. The serum NAbs and RBD-specific IgG antibodies GMTs were calculated with the two-side 95% confidence intervals (CIs) on the basis of the t-distribution of the log-transformed titers, and were then back-transformed to the original scale. Geometric mean fold increases (GMFIs) of NAbs and RBD-specific binding antibodies and GMT ratios of heterologous boost group versus homologous boost group were also calculated. Data below the detection limit were assigned a value half of that limit and data above the highest detection limit were assigned a value of the threshold. χ^2^ test and Fisher’s exact test were used to analyze categorical data when appropriate. Analysis of variance was used to analyze the log-transformed antibody titers and the Wilcoxon rank-sum test was used for non-normal distributed data. SAS (version 9.4) and GraphPad Prism (version 8.0.1) were used for statistical analyses.

## Results

### Baseline characteristics

Between September 2021 and March 2022, 418 (99.8%) participants completed the 3-month follow-up (140 [100.0%] in the low-dose group, 139 [100.0%] in the high dose group, 139 [99.3%] in the CoronaVac group). 118 (28.2%) participants donated blood samples (40 [28.6%] in the low-dose group, 38 [27.3%] in the high dose, 40 [28.6%] in the CoronaVac group) at month 3 post-boost. 413 (98.6%) participants completed the 6-month follow-up (139 [99.3%] in the low-dose group, 137 [98.6%] in the high dose group, 137 [97.9%] in the CoronaVac group), and 114 (27.2%) contributed blood samples (38 [27.1%] in the low-dose group, 37 [26.6%] in the high dose, 39 [27.9%] in the CoronaVac group) at month 6 post-boost for antibody persistence analysis (**Figure 1**). Participants in the antibody persistence analysis cohort had a mean age of 39 years with a standard deviation of 11 years and 68 (56.7%) were female. Of whom, the median time interval (interquartile range) between the second prime dose of CoronaVac and the booster was 5.0 (IQR 4.0, 5.0) months. Demographic characteristics of the participants were comparable cross the groups (**Supplementary Table 1**).

**Figure 1.**
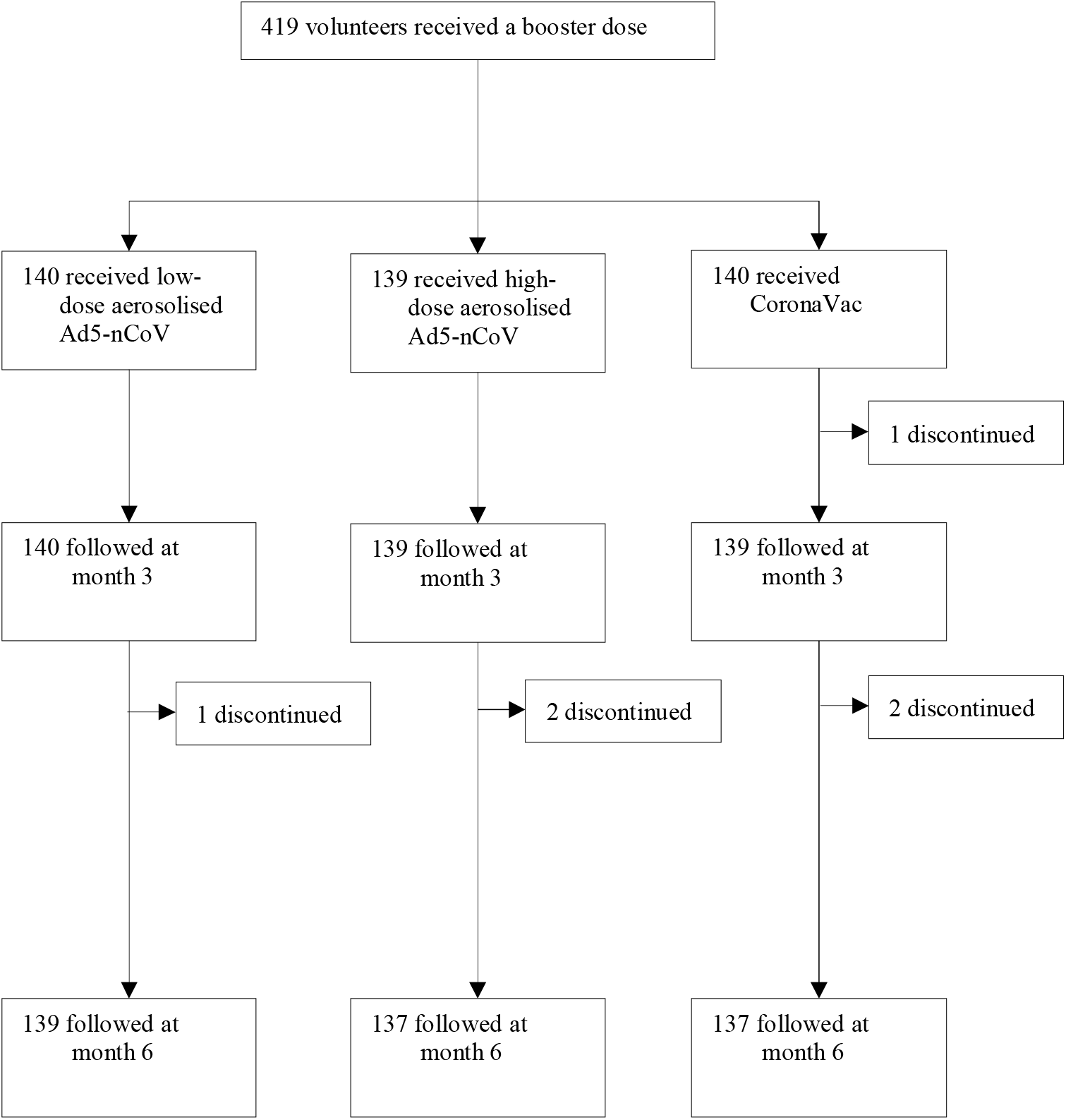
Study profile. Ad5-nCoV = orally administered aerosolised adenovirus type-5 vector-based COVID-19 vaccine carrying full-length SARS-CoV-2 spike gene. CoronaVac= intramuscularly administered inactivated whole virion SARS-CoV-2 vaccine.

### NAbs against wild-type SARS-CoV-2

At day 28 after the booster vaccination, NAb GMTs against live wild-type SARS-CoV-2 were 1937.3 (95% CI 1466.9, 2558.4) in the low-dose group, 1350.8 (95% CI 952.6, 1915.3) in the high-dose group and 73.5 (95% CI 52.3, 103.3) in the CoronaVac group. The NAb GMT at day 28 of the low-dose and high-dose aerosolised Ad5-nCoV groups were significantly higher than that of the CoronaVac group, with titers 26.4 folds and 18.4 folds higher compared with that in the CoronaVac group at day 28 post-boost (p<0.0001), respectively. At month 3, NAb GMTs against live wild-type SARS-CoV-2 decreased to 530.1 (95% CI 412.5, 681.1) in the low-dose group, 457.6 (95% CI 349.4, 599.2) in the high-dose group and 20.4 (95% CI 14.3, 29.1) in the CoronaVac group. However, the NAb GMT of low-dose and high-dose groups at month 3 were 26.0 folds and 22.4 folds higher than that of the CoronaVac group at month 3 post-boost (p<0.0001). At month 6, NAb GMTs against live wild-type SARS-CoV-2 further decreased to 312.9 (95% CI 237.7, 411.8) in the low-dose group, 251.1 (95% CI 178.2, 354.0) in the high-dose group and 10.4 (95% CI 7.8, 14.0) in the CoronaVac group. However, compared with the NAb GMT of CoronaVac group at month 6 post-boost, the NAb GMT of low-dose and high-dose groups were 30.1 folds and 24.1 folds higher, respectively (p<0.0001) (**Table 1, Figure 2**). There was no significant difference between the low-dose and high-dose aerosolised Ad5-nCoV groups at either day 28 (p=0.5265), month 3 (p=0.4206) or month 6 (p=0.3112) after the booster dose. In the low-dose and high-dose aerosolised Ad5-nCoV groups, the NAb GMTs decreased by 72.6% and 66.1% at month 3, 83.9% and 81.4% at month 6 after the booster dose, respectively, compared with 28 days post-boost. While, in the CoronaVac group, the NAb GMTs decreased by 72.2% and 85.9%, respectively (**Figure 2**).

**Table 1.** GMT, seropositivity rate, GMFI and GMT ratio of neutralizing antibodies to wild-type SARS-CoV-2 after a booster vaccination.

|  | Low-dose group<br>(n=40) | *P* value* | High-dose group<br>(n=40) | *P* value ** | CoronaVac group<br>(n=40) |
| --- | --- | --- | --- | --- | --- |
| Day 28 |  |  |  |  |  |
| GMT | 1937.3(1466.9, 2558.4) | <0.0001 | 1350.8(952.6, 1915.3) | <0.0001 | 73.5(52.3, 103.3) |
| Seropositivity | 100.0(91.2, 100.0) | 1.0000 | 100.0(90.5, 100.0) | 1.0000 | 97.5(86.8, 99.9) |
| GMFI | 519.1(357.7, 753.2) | <0.0001 | 323.9(216.1, 485.7) | <0.0001 | 17.9(12.3, 25.9) |
| GMT ratio | 26.4(17.1, 46.7) | — | 18.4(11.4, 29.7) | — | — |
| Month 3 |  |  |  |  |  |
| GMT | 530.1(412.5, 681.1) | <0.0001 | 457.6(349.4, 599.2) | <0.0001 | 20.4(14.3, 29.1) |
| Seropositivity | 100.0(91.2, 100.0) | 0.0255 | 100.0(90.5, 100.0) | 0.0260 | 85.0(70.2, 94.3) |
| GMFI | 139.6(100.7, 193.4) | <0.0001 | 110.2(76.7, 158.2) | <0.0001 | 5.6(3.6, 8.5) |
| GMT ratio | 26.0(18.2, 45.2) | — | 22.4(15.7, 38.9) | — | — |
| Month 6 |  |  |  |  |  |
| GMT | 312.9(237.7, 411.8) | <0.0001 | 251.1(178.2, 354.0) | <0.0001 | 10.4(7.8, 14.0) |
| Seropositivity | 100.0(90.8, 100.0) | <0.0001 | 100.0(90.3, 100.0) | <0.0001 | 66.7(49.8, 80.9) |
| GMFI | 84.1(58.2, 121.7) | <0.0001 | 60.4(40.0, 91.3) | <0.0001 | 2.8(2.0, 4.0) |
| GMT ratio | 30.1(18.8, 74.6) | — | 24.1(15.1, 59.5) | — | — |

Data are GMT (95% CI), seropositivity (%), 95% CI), GMT ratio (95% CI) or GMFI (95% CI). GMT=geometric mean titer. GMFI=geometric mean fold increase. Seropositivity (%): The proportion of participants whose antibody titers was defined as a detectable neutralizing antibody titer $\geq 1:8$ . \* The p values of this column are the results of comparison between low-dose aerosolised vaccine group and inactivated vaccine group. \*\* The p values of this column are the results of comparison between high-dose aerosolised vaccine group and inactivated vaccine group. GMT ratio=heterologous boost group/homologous boost group. Measurements on day 28 were taken 28 days after the booster vaccination.

**Figure 2.**
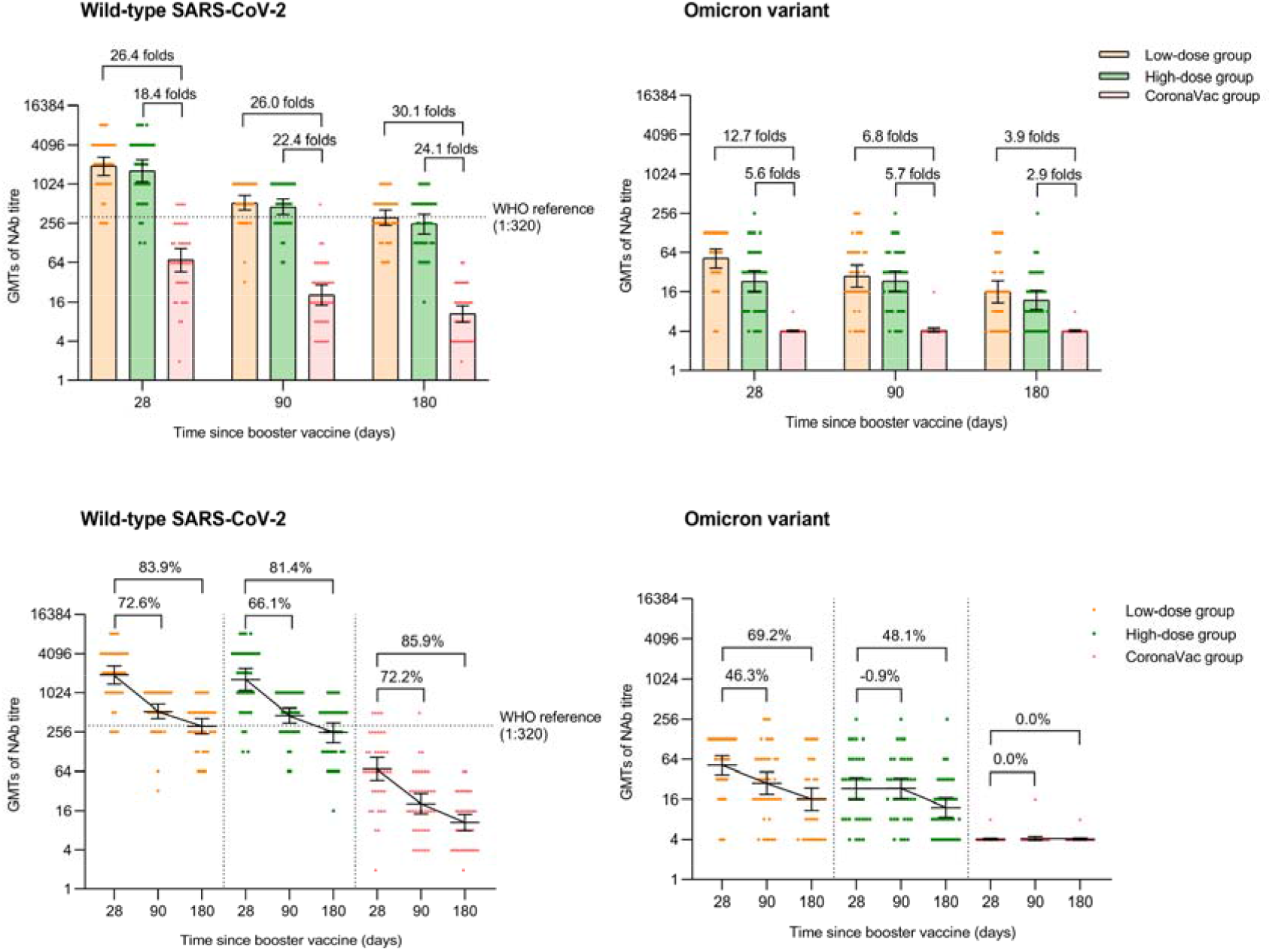
Neutralising antibodies against wild-type SARS-CoV-2 and Omicron variant after a booster vaccination. Horizontal bars show GMTs and error bars show 95% confidence intervals. Data (folds) above the bars are the NAb GMT ratios of the homologous boost group to the heterologous boost group. Long lines connecting the GMTs of adjacent groups indicate trends in neutralising antibodies over days after a booster vaccination. Data (%) above the bars show the percent reduction in GMTs of NAb titers at day 90 and day 180 post-boost compared to day 28. GMT=geometric mean titre in serum. NAb=neutralising antibody.

The low-dose and high-dose groups also showed higher NAb GMFIs against live wild-type SARS-CoV-2 than the CoronaVac group (519.1 [95% CI 357.7, 753.2] and 323.9 [95% CI 216.1, 485.7] versus 17.9 [95% CI 12.3, 25.9], respectively, p<0.0001) at day 28 post-booster. The NAb GMFIs decreased to 139.6 (95% CI 100.7, 193.4) in the low-dose group and 110.2 (95% CI 76.7, 158.2) in the high-dose group at month 3, and then to 84.1 (95% CI 58.2, 121.7) in the low-dose group and 60.4 (95% CI 40.0, 91.3) in the high-dose group, at month 6. While the NAb GMFIs of the CoronaVac group decreased to 5.6 (95% CI 3.6, 8.5) and 2.8 (95% CI 2.0, 4.0) at month 3 and 6. Additionally, all participants in the low-dose and high-dose groups had seropositivity at day 28 post-boost, while 39 (97.5%) participants in the CoronaVac had seropositivity. At month 3, all participants in the low-dose and high-dose groups had seropositivity, while 34 (85.0%) had seropositivity in the CoronaVac. At month 6, all participants in the low-dose and high-dose groups had seropositivity, while 26 (66.7%) participants in the CoronaVac had seropositivity (**Table 1**).

### NAbs against the omicron variant

At day 28 after the booster vaccination, NAb GMTs against live omicron variant B.1.1.529 were 52.0 (95% CI 37.2, 72.6) in the low-dose group and 23.1 (95% CI 15.7, 33.9) in the high-dose group. At month 3, NAb GMTs against live omicron variant B.1.1.529 were 27.9 (95% CI 18.8, 41.3) in the low-dose group and 23.3 (95% CI 16.2, 33.3) in the high-dose group. At month 6, NAb GMTs against live omicron variant B.1.1.529 decreased to 16.0 (95% CI 10.9, 23.5) in the low-dose group and to 12.0 (95% CI 8.5, 16.8) in the high-dose group. However, in the CoronaVac group, nearly all participants had no detective NAbs against omicron variant B.1.1.529 at any time point (**Table 2, Figure 2**). There was a significant difference between the two aerosolised Ad5-nCoV groups at day 28 (p=0.0018), and no significant difference between the two aerosolised Ad5-nCoV groups at either month 3 (p=0.4983) or month 6 (p=0.2593) after the booster dose. In the low-dose group, the NAb GMTs decreased by 46.3% and 69.2% at 3 and 6 months after the booster dose, respectively, compared with 28 days post-boost. In the high-dose group, the NAb GMT increased by 0.9% at month 3 and decreased by 48.1% at month 6 after the booster vaccination (**Figure 2**).

**Table 2:** GMT, seropositivity rate, and GMT ratio of neutralizing antibodies to omicron variant after a booster vaccination.

|  | Low-dose group<br>(n=40) | *P* value* | High-dose group<br>(n=37) | *P* value ** | CoronaVac group<br>(n=40) |
| --- | --- | --- | --- | --- | --- |
| Day 28 |  |  |  |  |  |
| GMT | 52.0(37.2, 72.6) | <0.0001 | 23.1(15.7, 33.9) | <0.0001 | 4.1(3.9, 4.2) |
| Seropositivity | 92.5(79.6, 98.4) | <0.0001 | 88.9(73.9, 96.9) | <0.0001 | 2.5(0.1, 13.2) |
| GMT ratio | 12.7(6.6, 178.6) | — | 5.6(3.0, 46.1) | — | — |
| Month 3 |  |  |  |  |  |
| GMT | 27.9(18.8, 41.3) | <0.0001 | 23.3(16.2, 33.3) | <0.0001 | 4.1(3.9, 4.4) |
| Seropositivity | 85.0(70.2, 94.3) | <0.0001 | 89.2(74.6, 97.0) | <0.0001 | 2.5(0.1, 13.2) |
| GMT ratio | 6.8(3.6, 64.1) | — | 5.7(3.0, 46.9) | — | — |
| Month 6 |  |  |  |  |  |
| GMT | 16.0(10.9, 23.5) | <0.0001 | 12.0(8.5, 16.8) | <0.0001 | 4.1(3.9, 4.2) |
| Seropositivity | 73.7(56.9, 86.6) | <0.0001 | 72.2(54.8, 85.8) | <0.0001 | 2.5(0.1, 13.5) |
| GMT ratio | 3.9(2.1, 23.6) | — | 2.9(1.6, 13.6) | — | — |

Data are GMT (95% CI), seropositivity (%), 95% CI, GMT ratio (95% CI). GMT=geometric mean titer. Seropositivity (%): The proportion of participants whose antibody titers was defined as a detectable neutralizing antibody titer $\geq 1:8$ . \*The *p* values of this column are the results of comparison between low-dose aerosolised vaccine group and inactivated vaccine group. \*\* The *p* values of this column are the results of comparison between high-dose aerosolised vaccine group and inactivated vaccine group. GMT ratio=heterologous boost group/homologous boost group. Measurements on day 28 were taken 28 days after the booster vaccination.

Moreover, at day 28, the seropositivity for NAbs against omicron variant B.1.1.529 in the low-dose and high-dose groups were 92.5% (95% CI 79.6, 98.4) and 88.9% (95% CI 73.9, 96.9), respectively. At month 3, the seropositivity was 85.0% (95% CI 70.2, 94.3) in the low-dose group and 89.2% (95% CI 74.6, 97.0) in the high-dose group. At month 6, the seropositivity decreased to 73.7% (95% CI 56.9, 86.6) in the low-dose group and to 72.2% (95% CI 54.8, 85.8) in the high-dose group. In the CoronaVac group, almost no participants were seropositive at any time point (**Table 2**). There were no significant differences in seropositivity for NAbs against omicron variant B.1.1.529 between the two aerosolised Ad5-nCoV groups at either day 28 (p=0.7015), month 3 (p=0.7385), or month 6 (p=0.8874).

### RBD-specific IgG antibodies

At day 28 after the booster vaccination, the GMTs of RBD-specific IgG antibodies were 5210.8 (95% CI 3797.8, 7149.5) in the low-dose group, 5743.4 (95% CI 4059.9, 8124.9) in the high-dose group and 294.2 (95% CI 223.7, 386.9) in the CoronaVac group. The GMTs of RBD-specific IgG antibodies of low-dose and high-dose groups were significantly higher than that of the CoronaVac group, with titres 17.7 folds and 19.5 folds higher compared with that in the CoronaVac group at day 28 post-boost, respectively (p<0.0001). At month 3, the GMTs of RBD-specific IgG antibodies were 3646.2 (95% CI 2707.0, 4911.3) in the low-dose group, 3328.5 (95% CI 2429.7, 4559.8) in the high-dose group and 138.8 (95% CI 102.9, 187.2) in the CoronaVac group. The GMTs of RBD-specific IgG antibodies of low-dose and high-dose groups were 26.2 folds and 24.0 folds higher compared with that in the CoronaVac group at month 3 post-boost, respectively (p<0.0001). At month 6, the GMTs of RBD-specific IgG antibodies were 2711.3 (95% CI 2040.9, 3601.8) in the low-dose group, 2218.2 (95% CI 1621.5, 3034.4) in the high-dose group and 74.7 (95% CI 53.0, 105.3) in the CoronaVac group. The GMTs of RBD-specific IgG antibodies of low-dose and high-dose groups were 36.2 folds and 29.7 folds higher than that in the CoronaVac group at month 6 post-boost, respectively (p<0.0001) (**Table 3, Figure 3**). There was no significant difference between the two aerosolised Ad5-nCoV groups at either day 28 (p=0.6762), month 3 (p=0.6712) or month 6 (p=0.3386) after the booster dose. In the low-dose group, the GMTs of RBD-specific IgG antibodies decreased by 30.0% and 48.0% at 3 and 6 months after the booster dose, respectively, compared with 28 days post-boost. In the high-dose group, the GMTs of RBD-specific IgG antibodies decreased by 42.1% at month 3 and by 61.4% at month 6 after the booster vaccination. In the CoronaVac group, the NAb GMTs decreased by 52.8% and 74.6%, respectively, at the same time points (**Figure 3**).

**Table 3:** GMT, GMFI and GMT ratio of RBD-specific IgG antibodies after a booster vaccination.

|  | Low-dose group<br>(n=40) | *P* value* | High-dose group<br>(n=40) | *P* value ** | CoronaVac group<br>(n=40) |
| --- | --- | --- | --- | --- | --- |
| Day 28 |  |  |  |  |  |
| GMT | 5210.8(3797.8, 7149.5) | <0.0001 | 5743.4(4059.9, 8124.9) | <0.0001 | 294.2(223.7, 386.9) |
| GMFI | 203.9(127.1, 327.2) | <0.0001 | 198.4(121.4, 324.2) | <0.0001 | 14.5(9.8, 21.3) |
| GMT ratio | 17.7(15.9, 19.9) | — | 19.5(17.6, 22.0) | — | — |
| Month 3 |  |  |  |  |  |
| GMT | 3646.2(2707.0, 4911.3) | <0.0001 | 3328.5(2429.7, 4559.8) | <0.0001 | 138.8(102.9, 187.2) |
| GMFI | 142.7(92.8, 219.4) | <0.0001 | 120.2(76.3, 189.4) | <0.0001 | 6.8(4.5, 10.3) |
| GMT ratio | 26.2(22.6, 31.3) | — | 24.0(20.6, 28.7) | — | — |
| Month 6 |  |  |  |  |  |
| GMT | 2711.3(2040.9, 3601.8) | <0.0001 | 2218.2(1621.5, 3034.4) | <0.0001 | 74.7(53.0, 105.3) |
| GMFI | 108.7(71.3, 165.8) | <0.0001 | 80.4(50.3, 128.4) | <0.0001 | 3.6(2.3, 5.6) |
| GMT ratio | 36.2(29.7, 46.7) | — | 29.7(24.3, 38.2) | — | — |

Data are GMT (95% CI), GMFI (95% CI) or GMT ratio (95%CI). GMT=geometric mean titer. GMFI=geometric mean fold increase. GMT ratio=heterologous boost group/homologous boost group. \* The p values of this column are the results of comparison between low-dose aerosol vaccine group and inactivated vaccine group. \*\* The p values of this column are the results of comparison between high-dose aerosol vaccine group and inactivated vaccine group. Measurements on day 28 were taken 28 days after the booster dose.

**Figure 3.**
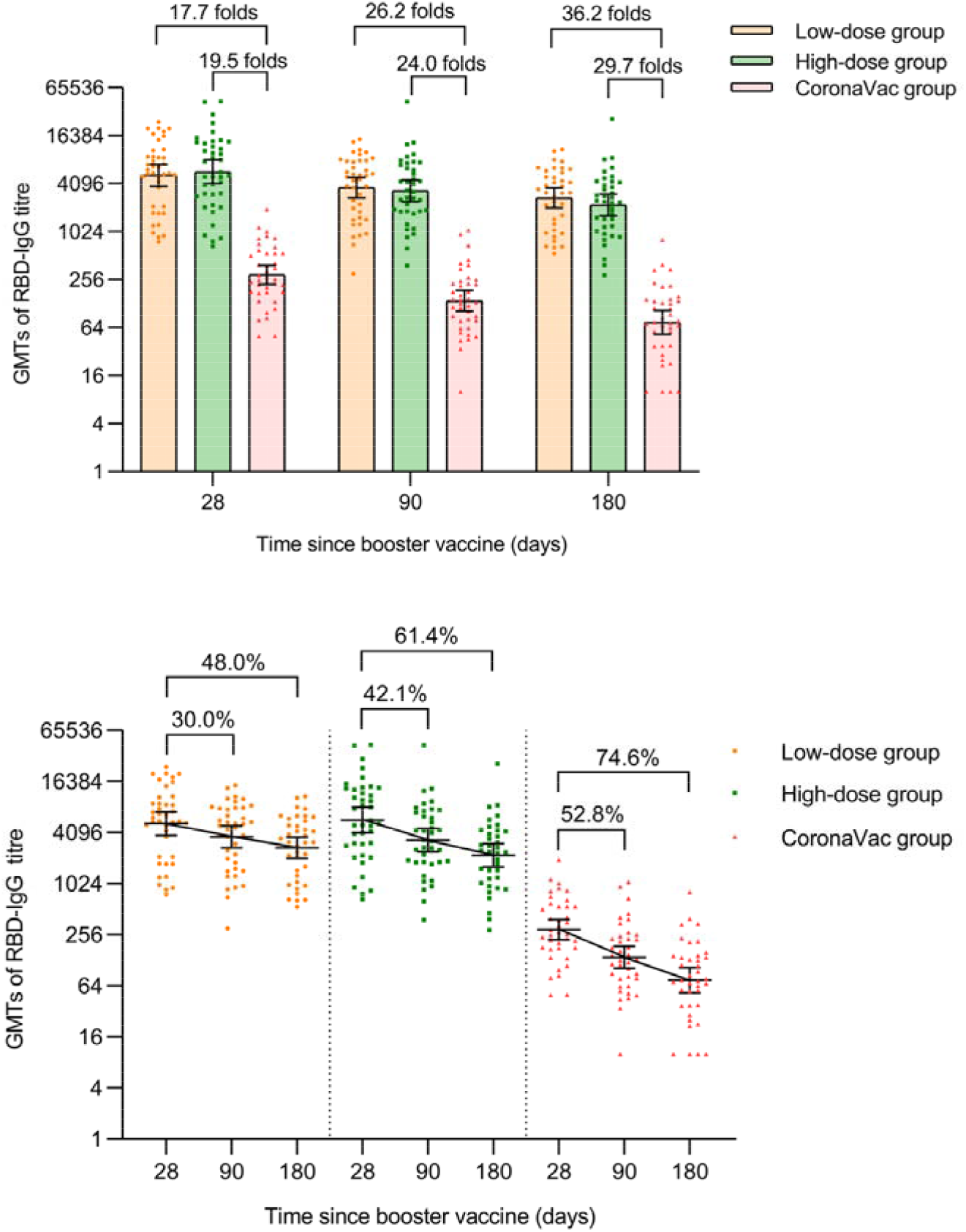
RBD-specific IgG antibodies after a booster vaccination. Horizontal bars show geometric mean titres and error bars show 95% confidence intervals. Data (folds) above the bars are the NAb GMT ratios of the homologous boost group to the heterologous boost group. Long lines connecting the geometric mean titers of adjacent groups indicate trends in RBD-IgG antibodies over days after a booster vaccination. Data (%) above the bars show the percent reduction in GMTs of RBD-IgG antibody titers at day 90 and day 180 post-boost compared to day 28. GMT=geometric mean titre in serum. RBD-IgG=receptor-binding domain (RBD)-specific IgG antibodies.

The low-dose and high-dose groups had higher GMFIs of RBD-specific IgG antibodies than the CoronaVac group (203.9 [95% CI 127.1, 327.2] and 198.4 [95% CI 121.4, 324.2] versus 14.5 [95% CI 9.8, 21.3], respectively, p<0.0001) at day 28 post-booster. The GMFIs of RBD-specific IgG antibodies decreased to 142.7 (95% CI 92.8, 219.4) in the low-dose group and 120.2 (95% CI 76.3, 189.4) in the high-dose group at month 3 post-boost, compared with 6.8 (95% CI 4.5, 10.3) in the CoronaVac group (p<0.0001). Then the GMFIs of RBD-specific IgG antibodies decreased to 108.7 (95% CI 71.3, 165.8) in the low-dose group and 80.4 (95% CI 50.3, 128.4) in the high-dose group, compared with 3.6 (95% CI 2.3, 5.6) in the CoronaVac group (p<0.0001) at month 6 (**Table 3**).

### Serious Adverse Events and pregnancy events

A total of three (2.5%) participants reported SAEs during the study period. Two (1.4%) occurred in the low-dose group, one of which was a 60-year-old man hospitalized at day 28 after the booster vaccine due to a 6-year history of pulmonary nodules; the other, a 46-year-old woman, was hospitalized at day 132 after a booster dose for a 20-year history of recurrent headaches. One (0.7%) occurred in the CoronaVac group, a 53-year-old male who was hospitalized at day 146 post-boost because of a 2-month history of cough and asthma. All three participants improved within 5 days after the hospitalization, and all these events were determined to be unrelated to vaccination (**Table 4**). In addition, four (3.3%) female participants became pregnant during the follow-up period: three (2.1%) in the low-dose group had the last menstrual period at day 117, day 38, and day 91 after the booster dose, respectively, and were 8 weeks, 21 weeks, and 14 weeks plus 1 day gestation at the 6th month follow-up time. One (0.7%) in the CoronaVac group had the last menstrual period at day 33 after the booster dose and was 21 weeks gestation at month 6. No adverse pregnancy outcomes and fetal abnormalities were observed during the 6-month follow-up after the booster vaccination (**Table 5**).

**Table 4.** Characteristics of serious adverse events during the study

| Subject ID | Group | Gender | Age | Time of onset (days after the booster vaccination) | Reason for hospitalization | Underling Disease | Days of hospital stay | Treatment | SAE Outcome | Relationship with vaccines |
| --- | --- | --- | --- | --- | --- | --- | --- | --- | --- | --- |
| V250 | Low-dose group | Male | 60 | 28 | Lung nodules for 6 years | 1. Lung nodules;<br>2. Hypothyroidism;<br>3. Hypertension | 4 | Right middle lobectomy | Improved | Irrelevant |
| V400 | Low-dose group | Female | 46 | 132 | Repeated headaches for more than 20 years | Central atrial septal defect (foramen ovale) | 4 | Percutaneous foramen ovale closure | Improved | Irrelevant |
| V405 | CoronaVac group | Male | 53 | 146 | Cough and asthma for more than 2 months | 1. Cardiac insufficiency<br>2. Coronary atherosclerotic heart disease<br>3. Bilateral pleural effusion<br>4. Atrial fibrillation<br>5. Pericardial effusion<br>6. Type 2 diabetes<br>7. Hypertension | 5 | Symptomatic treatment such as anti-infection, anticoagulation, ventricular rate control, and cardiac load reduction | Improved | Irrelevant |

**Table 5.** Characteristics of pregnancy events during the study

| Subject ID | Group | Age (years) | Height (cm) | Weight (kg) | Last menstrual period (days after the booster vaccination) | Length of pregnancy when reporting to investigators | Concomitant medication |
| --- | --- | --- | --- | --- | --- | --- | --- |
| V390 | Low-dose group | 30 | 158 | 48 | 117 | 8 weeks | None |
| V306 | Low-dose group | 32 | 160 | 50 | 38 | 21 weeks | None |
| V271 | Low-dose group | 24 | 166 | 50 | 91 | 14 weeks + 1 day | None |
| V085 | CoronaVac group | 31 | 172 | 60 | 33 | 21 weeks | None |

## Discussion

Previous reported study indicated that, in Chinese adults aged 18 years and older, heterologous aerosolized Ad5-nCoV plus two-dose CoronaVac was more immunogenic than three-dose CoronaVac within 28 days post-vaccination [11]. In this report, we found that the NAb GMTs against the wild-type SARS-CoV-2 and the omicron variant at month 6 after the heterologous aerosolized inhalation of 0.1 or 0.2 mL Ad5-CoV booster vaccine were higher than those at day 28 after the homologous CoronaVac booster vaccine, in adults who received two-dose CoronaVac. The superiority of the NAb responses to live wild-type SARS-CoV-2 and omicron variant in those receiving heterologous aerosolized Ad5-nCoV plus two-dose CoronaVac were persist for at least the 6-month follow-up period. NAb responses against the wild-type SARS-CoV-2 or the omicron variant waned over time from day 28 to month 6 after the third booster vaccine in all regimens. Similarly, low-dose and high-dose aerosolized booster vaccine recipients had substantially higher RBD-IgG antibody GMTs than CoronaVac booster vaccine recipients. RBD-IgG GMT was higher at 6 months after the heterologous regimen compared to that at day 28 after the homologous regimen. Moreover, we found no vaccine-related safety concerns in our cohort.

The immunogenicity of using homologous or heterologous vaccine as the third dose in adults who had received with two-dose CoronaVac at 3 months after the booster dose have been previously reported [12-15]. Compared to these studies, we presented data for longer follow-up period. Our study found that the NAb level at month 6 after the heterologous aerosolized Ad5-nCoV plus two-dose CoronaVac regimen was higher than the peak NAb level after three-dose CoronaVac (at day 28 after the booster dose). This result, reported for the first time, appeared to be more prominent than previously reported antibody levels following heterologous immunization.

Increasing antibody levels on day 28 after vaccination is critical for subsequent antibody persistence. As we found, in the homologous or heterologous booster groups, NAb levels for wild-type SARS-CoV-2 decreased by more than 80% at month 6 post-boost compared to day 28 post-boost. The rate of decline in antibody levels over time was similar in the three groups. The peak level of antibody in the heterologous booster group was higher than that in the homologous booster group. And antibody responses persisted longer in the heterologous booster group compared to the homologous booster group.

Orally administered aerosolised vaccines might become a valuable addition to the existing SARS-CoV-2 vaccine pools. Respiratory mucosal immunity was reported to potentially provide more protection by eliciting innate and adaptive immunity in the mucosal tissue at the site of virus entry, compared to intramuscular vaccines [16]. In a clinical trial, inhalation of two doses of live-attenuated influenza virus vector-based intranasal SARS-CoV-2 vaccine were administered as a primary immunization series on days 0 and 14 or on days 0 and 21 [17]. In another clinical trial, two-dose orally administered aerosolised Ad5-nCoV were performed on days 0 and 28 [18]. In both studies, a short time interval (2-4 weeks) between two-dose vaccination might have resulted in the partial neutralization of the second dose of vaccine by anti-viral vector antibodies produced following the first dose, because studies have been shown that the vaccine efficacy of adenoviral vector vaccine (ChAdOx1 nCoV-19, AZD-1222) tended to be higher when the time interval between the two intramuscular doses was more than 12 weeks [19, 20]. Another study showed that heterologous boosting with intramuscular AD5-nCOV was more immunogenic than homologous boosting with CoronaVac administered at an interval of 3–6 months after two doses of CoronaVac. The peak NAb GMT against SARS-CoV-2 after the heterologous booster was 616.9 IU ml^-1^ (95% CI 524.1, 726.3 IU ml^-1^) using the WHO international standard (Methods) [21]. Our study provided substantially greater results (a peak NAb GMT against SARS-CoV-2 of 6054.1 IU ml^-1^ [95% CI 4584.1, 7995.0 IU ml^-1^]) using orally administered aerosolized Ad5-nCoV as a heterologous booster vaccine after a median time of 5 months after two-dose CoronaVac compared to using intramuscular CoronaVac as a homologous booster vaccine. These findings suggest that a heterologous prime-boost regimen might avoid the negative effects of anti-viral vector antibodies.

Additionally, orally administered aerosolised vaccines could save vaccine doses compared to intramuscular vaccines on the premise of ensuring a non-inferior or even stronger immune response. One study showed that immune response elicited by a third intradermal dose of the adenoviral vector vaccine (AZD1222) was noninferior to the third dose of standard intramuscular CoronaVac in persons who had previously received two-dose CoronaVac, but the intradermal dose required only 20% of the intramuscular dose [22]. Likewise, volunteers in our study who inhaled only 0.1 mL (20% of the intramuscular dose) or 0.2 mL of aerosolized Ad5-nCoV produced significantly higher antibody responses than 0.5 mL of inactivated COVID-19 vaccine administered intramuscularly as a third dose. This is significantly beneficial for addressing the challenge that limited resource in vaccine distribution in many parts of the world need to be addressed to improve global access to life-saving vaccines.

Considerable escape of the omicron variant to antibody neutralization induced by vaccination has been reported [23]. Chen and colleagues found that neutralizing activity against the Omicron strain was reduced by 5.9-fold compared to the ancestral strain, at day 28 after the third dose of CoronaVac homologous booster [24]. Similarly, our results showed that nearly all participants had no detective NAbs against omicron at any time point after the homologous booster dose. However, using low-dose aerosolized Ad5-nCoV as a booster dose, the seropositivity for NAbs against omicron variant was 92.5% (95%CI 79.6%, 98.4%), 85.0% (95%CI 70.2%, 94.3%) and 73.7% (95%CI 56.9%, 86.6%) on day 28, month 3, and month 6, respectively. When using high-dose aerosolized Ad5-nCoV as a booster dose, the seropositivity was similar to that in the low-dose aerosolized Ad5-nCoV group. Consistent with previous studies [25, 26], our study suggested that although omicron variant evaded NAb responses elicited by the homologous CoronaVac boost regimen, the heterologous boost regimen may provide protection against omicron variant.

This study had several limitations. Firstly, cellular and mucosal immunity data were not provided. Airway-secreted IgA elicited by mucosal immunity and resident memory B and T cells in the airway mucosa may provide an effective barrier to blocking viral invasion [16]. Secondly, vaccine efficacy or vaccine effectiveness against the original SARS-CoV-2 strain or omicron variants were not presented, although neutralizing antibodies could serve as surrogate markers of vaccine efficacy [27]. Thirdly, we analyzed only the original Omicron virus BA.1 for NAbs against live omicron variant, while NAbs against BA.2, or BA.4/5 subvariants were not detected in the laboratory. The BA.4 and BA.5 variants are more infectious than the original Omicron virus and substantially escape antibodies elicited by vaccination and BA.1 or BA.2 subvariant of omicron infection [28-30]. Therefore, this deserves more attention in future research. Finally, the influencing factors of antibody persistence had not been analyzed due to the small sample size. Previous studies found that prior infection and age influence the magnitude and persistence of antibody responses [31-33]. Immune memory appears to persist for at least 11 months in previously SARS-CoV-2 infected individuals. Future studies determining the antibody persistence of heterologous aerosolized Ad5-nCoV regimens in different age groups and special populations will be required.

This study implied that policymakers could use aerosolized Ad5-nCoV vaccine as a third booster for persons who have received two-dose CoronaVac. Besides enhancing antibody responses, orally administered aerosolised vaccines’ potential advantages over injected vaccines include ease of administration, perhaps even self-administration, which may speed up the vaccination process. Disposal of sharps after mass vaccination campaigns was avoided by not using needles and syringes. Each aerosolized Ad5-nCoV vaccination uses only one-fifth intramuscular injection dose, which has the advantage of maximize allocation of vaccine resources and being economically feasible in terms of cost-efficiency. In conclusion, our study demonstrated that heterologous orally aerosolised Ad5-nCoV plus two-dose Coronavac had a good long-term safety profile and was persistently more immunogenic than three-dose CoronaVac, providing additional support for using a mix-and-match approach.

## Data Availability

The results supporting the findings in this study are available upon request from the corresponding authors.

