## Supplementary Table 1 for "Antibody Persistence and Safety through 6 Months after Heterologous Orally Aerosolised Ad5-nCoV in individuals primed with two-dose CoronaVac previously"

Table S1. Demographic characteristics of the participants in this study.

|  | Low-dose group (n=40) | High-dose group (n=40) | CoronaVac group (n=40) |
| --- | --- | --- | --- |
| Gender |  |  |  |
| Male, n (%) | 12(42.5) | 20(50.0) | 15(37.5) |
| Female, n (%) | 23(57.5) | 20(50.0) | 25(62.5) |
| Age, years |  |  |  |
| 18-59, n (%) | 38(95.0) | 39(97.5) | 38(95.0) |
| ≥60, n (%) | 2(5.0) | 1(2.5) | 2(5.0) |
| mean (SD) | 38.1(11.9) | 39.8(10.0) | 40.1(11.5) |
| Months since the last prime dose of CoronaVac, median (IQR) | 5.0(4.0, 5.0) | 5.0(5.0, 5.0) | 5.0(5.0, 5.0) |

Notes: SD= standard deviation. IQR=Interquartile range.
